# TCR repertoire profiling revealed antigen-driven CD8+ T cell clonal groups shared in synovial fluid of patients with spondyloarthritis

**DOI:** 10.1101/2022.05.06.22274633

**Authors:** Ekaterina A. Komech, Anastasia D. Koltakova, Anna A. Barinova, Anastasia A. Minervina, Maria A. Salnikova, Evgenia I. Shmidt, Tatiana V. Korotaeva, Elena Y. Loginova, Shandor F. Erdes, Ekaterina A. Bogdanova, Mikhail Shugay, Sergey Lukyanov, Yury B. Lebedev, Ivan V. Zvyagin

## Abstract

Spondyloarthritis (SpA) comprises a number of inflammatory rheumatic diseases with overlapping clinical manifestations. Strong association with several HLA-I alleles and T cell infiltration into an inflamed joint suggest involvement of T cells in SpA pathogenesis. In this study, we performed high-throughput T cell repertoire profiling of synovial fluid (SF) and peripheral blood (PB) samples collected from a large cohort of SpA patients. We showed that synovial fluid is enriched with expanded T cell clones that are shared between patients with similar HLA genotypes and persist during recurrent synovitis. Using the recently published algorithm we discovered antigen-driven CD8+ clonal groups associated with risk HLA-B*27 or HLA-B*38 alleles. These clonal groups were enriched in SF and had higher frequency in PB of SpA patients vs healthy donors, suggesting their relevance to joint inflammation. Several of the identified groups were shared among patients with ankylosing spondylitis and psoriatic arthritis, suggesting existence of a common immunopathological mechanism of the diseases. In summary, our results provide supporting evidence for the role of antigen-driven CD8+ T cell clones in pathogenesis of SpA.

## INTRODUCTION

Spondyloarthritis (SpA) comprises several inflammatory rheumatic diseases with overlapping clinical manifestations, including ankylosing spondylitis (AS), psoriatic arthritis (PsA), reactive arthritis, enteropathic SpA, and undifferentiated SpA. Risk of SpA is associated with genetic factors related to the antigen-presenting system (e.g. B*27, B*38, B*39 alleles of HLA-I, and ERAP1/2) and immune response regulation (e.g. IL23R, IL6R, STAT3) (1). Considering accumulation of T lymphocytes in inflamed joints (2), this suggests the involvement of T cells in pathogenesis of SpA.

T cell receptor (TCR) repertoire profiling is a powerful methodology to study T cells in health and disease and to identify condition-associated T cell clones (3). Recently we and others discovered a specific TCRbeta motif in peripheral blood (PB) of HLA-B*27+ AS patients (4–6). This motif was reported in synovial fluid (SF) of HLA-B*27+ patients with reactive arthritis (7), and further confirmed in SF of AS patients (4). A number of studies also demonstrated the presence of different clonal T cell expansions in inflamed joints of PsA patients (8–10). Investigation of T cell repertoire of an inflamed joint provides insight into the role of T cells in local inflammation and allows identification of the disease-associated clonal expansions. However, previous studies in this area were limited either by the cohort size or by repertoire sequencing depth.

Here we performed high-throughput TCRbeta repertoire profiling of SF collected once or twice from a total of 28 patients with AS or PsA and identified on average 28,000 distinct clonotypes per sample. We analysed general characteristics of SF clonal repertoire, its diversity, sharing and convergence, and investigated presence of specific T cell clonal expansions restricted to SpA-associated HLA-I alleles.

## RESULTS

### Synovial fluid of SpA patients contains expanded T cell clonotypes shared among donors with similar HLA genotypes

To study the TCRbeta repertoire we collected PB and SF samples from 10 AS and 17 PsA patients. The samples included total MNCs, as well as CD4+ and CD8+ T cell subsets isolated from at least 3×10^6 mononuclear cells. We also included repertoires of 25 AS patients sequenced in previous work to the analysis (4), thus, collectively analysing SF and PB samples from 28 and 51 donors, respectively (**Supplementary Table 1**). All TCRbeta repertoires were reconstructed using the protocol that implements double barcoding of samples and labelling of cDNA with unique molecular identifiers. The median number of unique TCRbeta cDNA molecules identified per sample was 267,258 for PB (IQR 144,492 - 483,323) and 207,539 for SF (IQR 83,554 - 358,229), reflecting comparable repertoire analysis depth of on average ∼250,000 T cells (**Supplementary Table 2**) (11).

First, we investigated the functional state of T cells in an inflamed site by evaluating the expression of PD-1 and CD137, which are induced upon TCR-dependent activation. On average ∼80% of SF T cells expressed PD-1 and 3-5% T cells expressed CD137 in contrast to less than 10% and 0.5% in PB, respectively (**Supplementary Figure 1**). Thus, SF T cells show signs of recent activation by cognate antigens.

To characterise the structure of SF repertoire we compared the clonal diversity and clonal size distribution between SF and PB samples. We also assessed the uniqueness of SF repertoire by estimating the PB-SF repertoire overlap. As the diversity metrics and clonal sharing are extremely affected by sample size (12), we equalised the repertoire analysis depth by sampling the 34,000 unique TCRbeta cDNAs from each dataset. Both CD8+ and CD4+ SF repertoires had significantly lower clonal diversity and larger clonal expansions compared with those of PB (Figure 1A-B). On average only ∼6% of PB clonotypes, defined by nucleotide sequence, were present in matched SF sample independent of CD4 or CD8 lineage.

**Figure 1.**
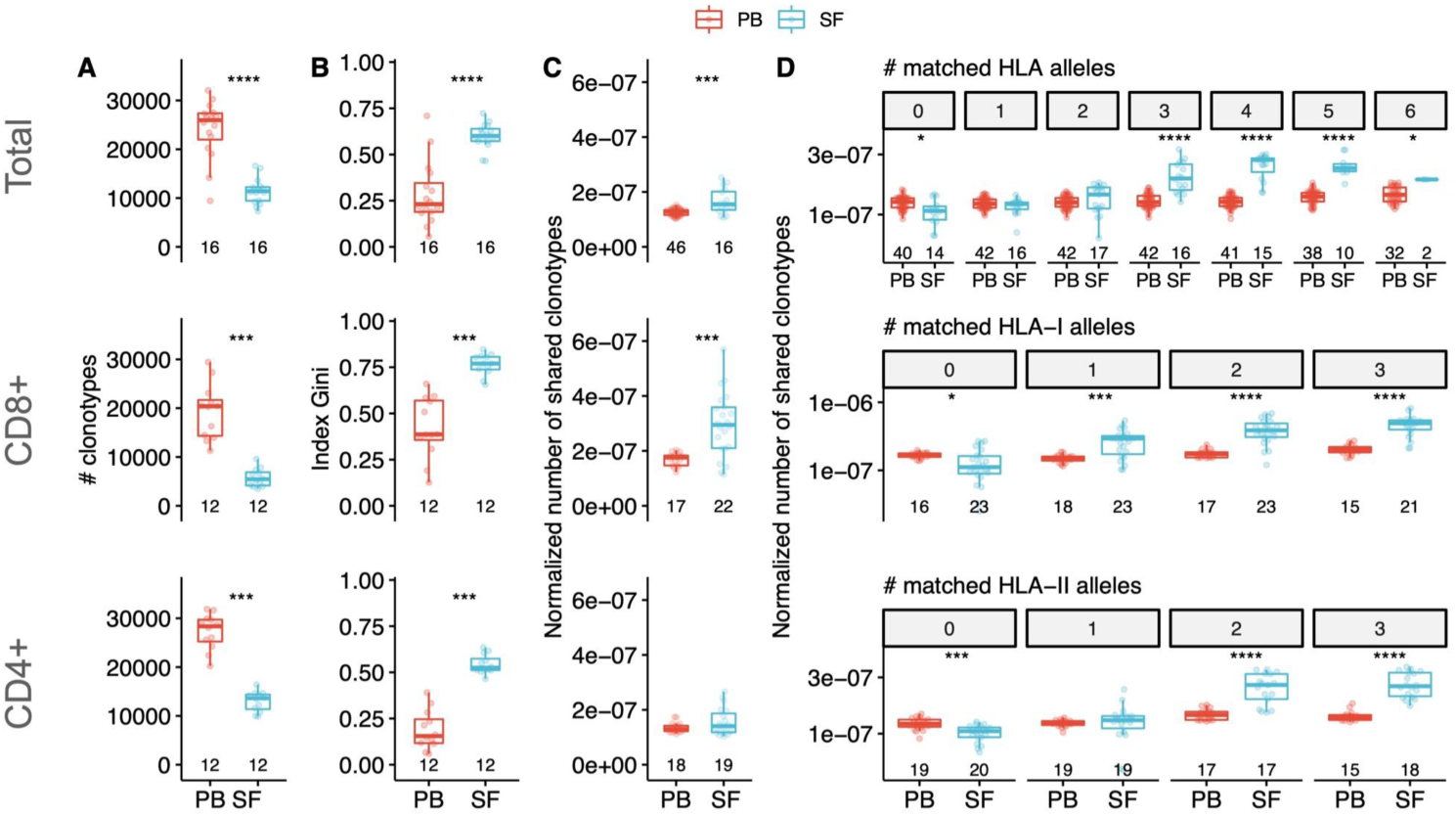
Characteristics of clonal T cell repertoire of synovial fluid from patients with SpA. Comparison of clonal diversity (**A**) and oligoclonality (**B**) between PB and SF repertoires of SpA patients (by two-sided Wilcoxon signed-rank test). **C, D** Normalised number of shared clonotypes per donor in total (**D**) or depending on the number of matched HLA alleles (**E**) between donors (by two-sided Wilcoxon’s rank sum test with FDR adjustment). Values below the box plots denote the number of donors. Top, middle and bottom panels correspond to the repertoires of total, CD8+ and CD4+ T cells, respectively. PB - peripheral blood, SF - synovial fluid, HLA - human leukocyte antigen. * p<0.05, ** p<0.01, *** p<0.001, **** p<0.0001

The total and CD8+ SF repertoires had higher clonal sharing (i.e. number of identical amino acid TCRbeta sequences) between individuals compared with those of PB (**Figure 1C**). Moreover, clonal sharing depended on the number of HLA alleles matched between donors: SF repertoires shared more clonotypes compared with PB only if donors shared at least one HLA-I allele (for CD8+ T cells) or two HLA-II alleles (for CD4+ T cells) (**Figure 1D**). Collectively, these data demonstrate that SF of SpA patients has a distinct structure of clonal repertoire and contains a specific set of expanded T cell clones that have the same antigen-specificity and are shared between individuals with similar HLA genotypes.

### Identification of clonotypes involved in immune response in synovial fluid

To search for the putative disease-associated T cell clonal groups we employed the ALICE algorithm that identifies co-presence of highly similar clonotypes typically arising during an immune response to the same antigen (13,14).

The ALICE was applied to each of the 28 CD8+ SF repertoires yielding on average ∼300 significant, i.e. having more homologs than expected by chance, amino acid clonotypes per sample (median 192, IQR 27-343). We then combined all those clonotypes (n=4432) from all samples and clustered them into similarity networks based on the identity of V-J-segments and CDR3 length with 1 amino acid substitution (13–17). Overall we got 1449 clusters: 1377 clusters were present in a single patient, while the others were shared, and 29 of them were shared by at least 3 patients (**Figure 2A**).

**Figure 2.**
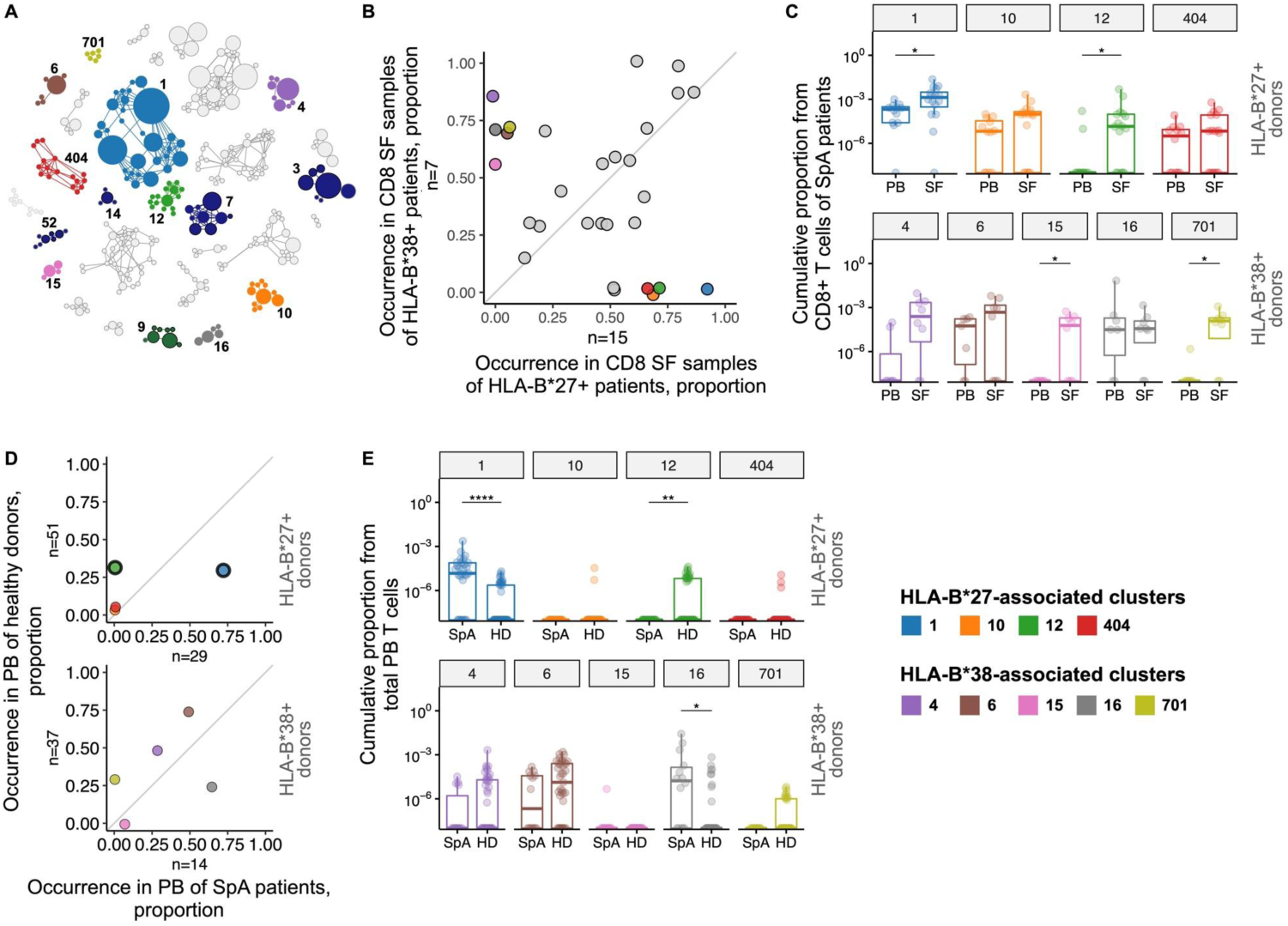
T cell clonotypes involved in immune response in synovial fluid of SpA patients. **A**, Similarity network of SF CD8+ clonotypes identified by the ALICE. Only 29 clusters shared by ≥3 patients are shown. Each node represents a unique amino acid clonotype. Edges connect nodes with one mismatch. Node’s size corresponds to the number of donors sharing this clonotype. Colour highlights clusters associated with HLA-B*27, HLA-B*38, or found in VDJdb. **B**, Occurrence of the clusters in CD8+ SF samples of HLA-B*27+ vs HLA-B*38+ patients. **C**, T cell frequency of HLA-B*27-associated (upper, n(PB)=12, n(SF)=16) or HLA-B*38-associated (bottom, n(PB)=7, n(SF)=8) clusters in CD8+ repertoires of patients. Occurrence (**D**) and T cell frequency (**E**) of HLA-B*27-associated (upper, n(SpA)=29, n(HD)=51) or HLA-B*38-associated (bottom, n(SpA)=14, n(HD)=37) clusters in total PB repertoires of SpA patients and HD. Black circles mark significantly enriched clusters (Fisher’s exact test with FDR-adjusted p<0.05). PB - peripheral blood, SF - synovial fluid, SpA - spondyloarthritis, HD - healthy donors. Two-sided Wilcoxon’s rank sum test with FDR adjustment * p<0.05, ** p<0.01, **** p<0.0001

Using VDJdb, a database of TCRs with known specificity (15), we found that 5 of the 29 shared clusters comprised clonotypes associated with HLA-A*02-restricted epitopes from EBV (clusters 3, 7, 14 and 52) and Influenza A virus (cluster 9). Accordingly, these clusters were detected exclusively in HLA-A*02+ individuals. Notably, the EBV-associated clonotypes were enriched in SF compared with PB samples (**Supplementary Figure 2**).

We further focused on the association of the identified clusters with two SpA risk HLA-I alleles, HLA-B*27:05 and HLA-B*38:01 (present as a HLA-B*38:01/C*12:03 haplotype) that were present in almost mutually exclusive manner in our cohort (n=16 and n=8, respectively, including one HLA-B*27+/B*38+ donor). Nine clusters were significantly associated with the presence of the alleles: clusters 1, 10, 12, 404 were overrepresented in CD8+ SF repertoires of HLA-B*27+ patients, and clusters 4, 6, 15, 16, 701 were overrepresented in repertoires of HLA-B*38+ patients (**Figure 2B** and **Supplementary Table 3**).

Clonotypes from cluster 1 matched or were similar to previously identified AS-related TCRbeta motif (4–6) and were detected in 15 out of 16 CD8+ SF samples of HLA-B*27+ patients, including HLA-B*27+/B*38+ donor (**Supplementary Table 3**). The most shared clonotype of cluster 4 TRBV10-2_CASSESPGNSNQPQHF_TRBJ1-5 was reported to be HLA-C*12:03-restricted and accordingly present in CD8 SF samples of 6 out of 8 HLA-B*38:01+/C*12:03+ patients and 1 out of 3 B*38:01-/C*12:03+ patients (18).

To address the relevance of the identified clusters to the joint inflammation we analysed their enrichment in SF, as well as the prevalence in PB repertoires of SpA patients in comparison with a large cohort of healthy donors (HD, from previously published studies) (4,19). The T cell frequency (i.e. cumulative proportion of T cells occupied by clonotypes of a cluster) of clusters 1, 12, 15 and 701 was significantly higher in CD8+ SF than in PB repertoires of HLA-B*27+ or HLA-B*38+ patients (**Figure 2C, Supplementary Figure 2**). In total PB samples, only cluster 1 was significantly overrepresented and had higher T cell frequency in SpA patients compared with HD (**Figure 2D-E, Supplementary Table 4**). On the contrary, cluster 12 was barely present in blood of patients but had significantly higher incidence and T cell frequency in HD. Clusters 15 and 701 were also almost absent from PB of SpA patients and had zero (cluster 15) or low T cell frequency (cluster 701) in PB of HD. HLA-B*38-associated cluster 16 had higher T cell frequency in PB of patients vs HD, but it was not enriched in CD8+ SF compared with blood repertoires of patients. The T cell frequency of the other clusters was either extremely low (clusters 10 and 404, HLA-B*27-associated), or did not differed (clusters 4 and 6, HLA-B*38-associated) between total PB samples of HD and SpA patients, and these clusters were not significantly enriched in CD8+ SF samples.

### Presence of the same expanded clonotypes in an inflamed joint during recurrent synovitis

To study how SF clonal T cell repertoire varies between relapses of synovitis we collected repeated SF samples from 4 SpA patients (9-23 months apart).

The overall similarity of the repertoires at those time points was estimated with Morisita’s overlap index that accounts for both incidence and T cell frequency of identical clonotypes between samples. The index is bounded between 0 (completely different repertoires) and 1 (absolutely identical repertoires). SF repertoires of the same patient appeared highly similar for all donors (Morisita’s index median 0.759, IQR 0.639 - 0.827, compared with <1*10^-5 for unrelated samples). Furthermore, out of the 1000 most abundant clonotypes at time point 1 more than 800 (median 838.5, IQR 831.5 - 853.2) were also detected at time point 2, and more than 400 (median 464.5, IQR 425.5 - 521.0) remained among the top 1000 clonotypes, indicating the stability of major clonal expansions of SF.

By employing the ALICE algorithm we found that only ∼30% of inferred antigen-driven clonal expansions were novel at time point 2 (median 29.7% IQR 27.2% - 31.9%), while the majority of them were shared between the time points. Of the previously identified HLA-B*27-associated clusters the clonotypes of cluster 1 were observed in repeated SF samples of all 3 HLA-B*27+ donors, and clonotypes of clusters 12 and 404 persisted in one of the three donors.

## DISCUSSION

Here we applied high-throughput TCR repertoire profiling to the synovial fluid of a large cohort of SpA patients to characterise the repertoire structure and to identify T cell clones involved in disease pathogenesis. We demonstrated that both CD4+ and CD8+ SF clonal repertoires have low diversity and contain large clonal expansions distinct from those of blood, confirming and extending earlier observations (8–10,20). The cytometry data show that the majority of SF T cells exhibit signs of TCR-dependent activation in concordance with results obtained with single-cell RNAseq (20). The same clonotypes were present in repeated SF samples collected several months apart, suggesting that they persist in the joint as tissue-resident T cells. T cells with resident memory phenotype were recently discovered in SF of PsA patients (20,21).

We further demonstrated that identical clonotypes accumulate in SF of unrelated patients with similar HLA genotypes, suggesting convergent selection to the same antigens. Considering that the immune response to a single antigen:MHC complex is mediated by T cell clones with highly similar TCRs (13–17), we discovered clusters of similar clonotypes shared between CD8+ SF samples of patients. Nine of them were associated with SpA risk HLA-I alleles HLA-B*27 or HLA-B*38. Several clusters were detected in PD-1+ and/or CD137+ subsets of SF T cells, suggesting recent antigen-driven activation. Clonotypes belonging to the clusters 1 and 6, were also detected in tissue-resident memory T cell subset (CD3+CD103+CD69±) further suggesting their involvevement in local immune response (summarised in **Figure 3, Supplementary Table 5**).

**Figure 3.**
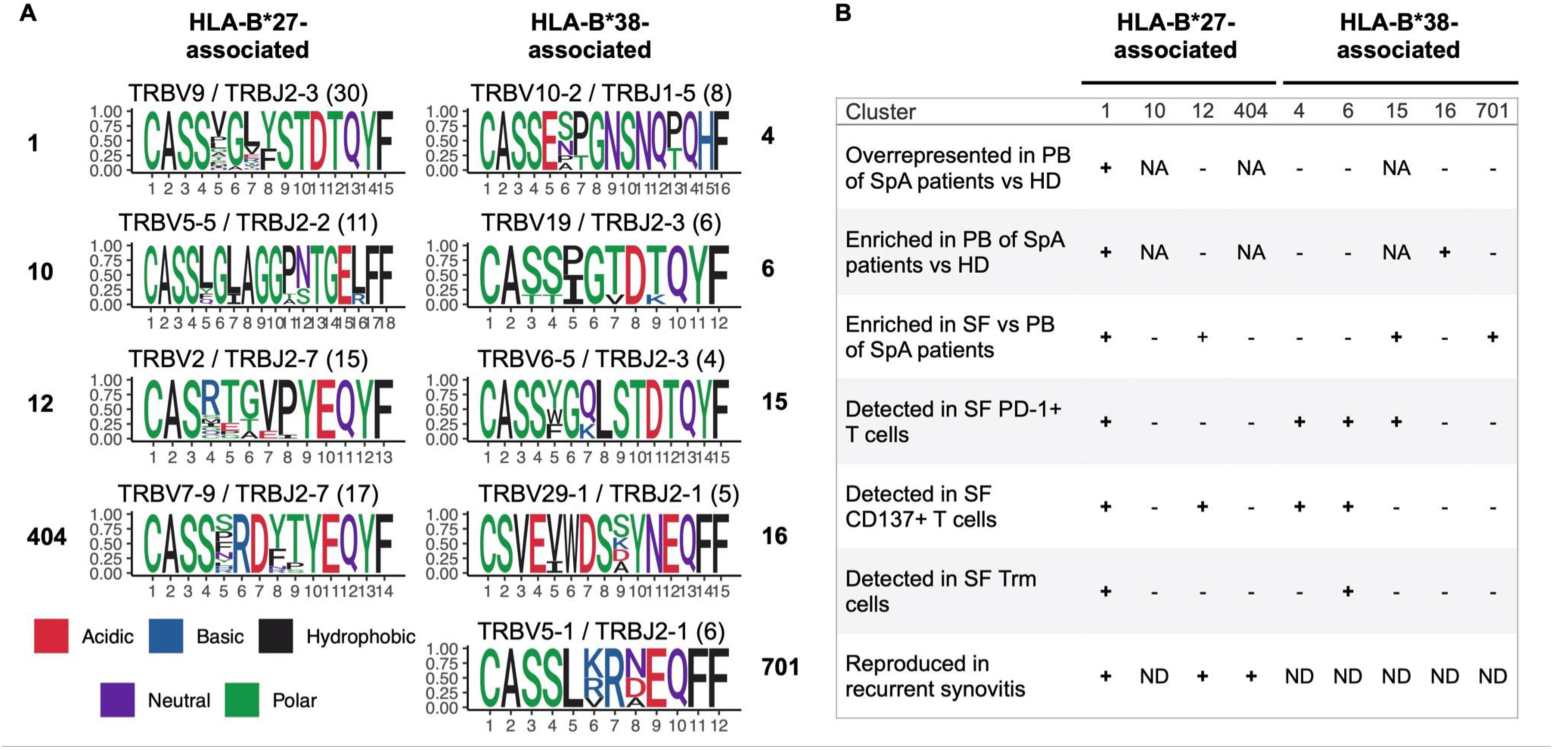
Characteristics of identified TCRbeta clusters. Sequence logo of CDR3 regions **(A**) and details (**B**) of the 9 identified clusters associated with HLA-B*27 or HLA-B*38. Values in parentheses represent the number of distinct clonotypes. PB - peripheral blood, SF - synovial fluid, Trm - tissue-resident memory T cells, NA - not applicable, ND - no data.

One of the HLA-B*27-associated clusters, cluster 1, was detected in SF of ∼94% HLA-B*27+ patients and matched TCRbeta motif previously identified in HLA-B*27+ AS and ReA patients (4–7). Here we report the presence of the same motif in SF of HLA-B*27+ patients with PsA, providing evidence for a common immunopathological mechanism of the diseases. We also identified three new TCRbeta clusters enriched in SF of HLA-B*27+ AS and PsA patients, and thus supposedly involved in the pathogenesis of SpA.

In SF of HLA-B*27- patients we discovered several TCRbeta motifs associated with HLA-B*38/C*12 haplotype. HLA-B*38-associated clusters behaved differently: three clusters demonstrated high (clusters 15 and 701) or moderate (cluster 4) enrichment in SF (**Figure 2C, Supplementary Figure 2**), and the two other clusters were represented comparably between PB and joint.

In agreement with the previous report (22), we found shared clusters assigned to HLA-A*02-restricted epitopes of EBV, and, cumulatively, the EBV-associated clonotypes were enriched in SF (**Supplementary Figure 3**). Note that it is common for virus-specific clones to be associated with T cell infiltration into inflamed sites, e.g. tumour-infiltrating T cells (23). These results draw attention to the potential role of virus-specific T cells in immune response to non-viral antigens.

Repertoire analysis based on TCRbeta sequence similarity and sharing between donors allowed us to identify clonal groups expanded in antigen-dependent manner in comparison with diverse bystander T cell clones (24). However, the association of a TCRbeta clonotype with a cluster could be inaccurate due to unknown paired TCRalpha chain and/or unclear impact of particular amino acid mismatches to the antigen-specificity. This, as well as potential heterogeneity of T cell phenotypes within a clone and/or cluster (reported recently for SF of PsA patients) (20) could affect the frequency and detection of a clonotype in different donors. It can be suggested that the dissimilarity in paired TCRalpha chain or T cell phenotype are related to the higher frequency of cluster 12 in PB of HD compared with SpA patients (**Figure 2C-D, Supplementary Figure 2**). Further studies designed to characterise paired TCRalpha repertoire, functional phenotype and antigen-specificity of the T cells belonging to the identified TCRbeta clusters are necessary to reveal their role in SpA pathogenesis.

In summary, we demonstrated that SF of SpA patients contains a distinct set of expanded T cell clones that persist over time, and discovered several TCRbeta motifs associated with risk HLA-I alleles HLA-B*27 and HLA-B*38. Our findings provide evidence for a role of antigen-driven CD8+ T cells in pathogenesis of SpA.

## MATERIALS AND METHODS

### Patients and samples

The study was conducted in accordance with the Declaration of Helsinki and approved by the local ethics committee of Pirogov Russian National Research Medical University. All patients gave their written informed consent for participation. We collected 28 SF and 51 PB samples from patients with PsA (n=17) or AS (n=35; T cell repertoires of 25 AS patients were taken from previously published study (4)). Full details on the cohort are provided in **Supplementary Table 1**. Patients were classified with ankylosing spondylitis or psoriatic arthritis according to modified New-York criteria or CASPAR, respectively (25,26). We also used previously published repertoires of healthy donors (4,19).

### Isolation of T cell subsets

Mononuclear cells (MNCs) from PB and SF samples were isolated by Ficoll density gradient centrifugation (PanEco). CD4+ and CD8+ T cells were isolated immediately from MNCs with anti-CD4 or anti-CD8 positive-selection magnetic beads (Dynabeads, Thermo Fisher Scientific). CD137+ and PD-1+ T cells, and CD103+CD69± T cells (tissue-resident memory) were sorted from cryopreserved SF MNCs using the following antibodies: CD3-PE/Cy5 (Beckman Coulter Cat# IM2635U, RRID:AB_10645166), CD8-PE/Cy5 (Beckman Coulter Cat# IM2638U, RRID:AB_131157), CD3-eFluor 450 (Thermo Fisher Scientific Cat# 48-0038-80, RRID:AB_1518801), PD-1-Brilliant Violet 421 (BioLegend Cat# 329920, RRID:AB_10960742), CD137-PE (Miltenyi Biotec Cat# 130-110-900, RRID:AB_2654985), CD103-FITC (Thermo Fisher Scientific Cat# 11-1031-81, RRID:AB_465175), CD69-PE/Cy5 (BioLegend Cat# 310907, RRID:AB_314842). Cell sorting was performed on FACSAria III (BD) directly into the RLT lysis buffer (Qiagen).

### TCRbeta library preparation, HLA-typing, and sequencing

Libraries of TCRbeta chains were prepared with the protocol that implements double barcoding of samples and labelling of cDNA with unique molecular identifiers, allowing for potential cross-sample contamination removal, error-correction, and data normalisation (described in (27)). In brief, total RNA from MNCs was isolated using TRIzol reagent (Thermo Fisher Scientific) or RNeasy Mini kit (Qiagen). Total RNA was used for cDNA synthesis using 5’RACE template switch technology to introduce universal primer binding site, unique molecular identifiers (UMI) and first sample barcode at the 5’ end of RNA molecules. Primers complementary to TCRbeta constant segments were used for initiation of cDNA synthesis. cDNA was amplified in two subsequent PCR steps. During the second PCR step, second sample barcode and adapters were introduced to the libraries.

The libraries were sequenced on NextSeq, HiSeq2000/2500 or MiSeq (Illumina) with 2×100bp or 2×150bp sequencing length.

HLA-genotyping was performed using an in-house NGS-based protocol.

### Data analysis and statistics

#### Raw data preprocessing

Raw sequencing reads were demultiplexed and clustered by UMI using MIGEC software (RRID:SCR_016337, version 1.2.9) with overseq threshold of 2 reads per UMI to exclude erroneous sequences (28). To generate the clonesets MiXCR software (RRID:SCR_018725, version 3.0.9) was used with settings for 5’RACE paired-end libraries (29). TCR repertoire analysis was performed using tcR R-package and custom R-scripts (30,31).

#### Repertoire diversity and clonal sharing

To assess the diversity metrics and clonal sharing all repertoires were normalised by sampling of 34,000 UMIs. Oligoclonality was measured by the Gini index that equals 0 in the even distribution of clonal frequencies and 1 in the monoclonal sample. Clonal sharing was calculated as the median of intersections (i.e. normalised number of identical amino acid TCRbeta sequences) per donor. Only paired PB and SF samples were included in the analysis. A two-sided Wilcoxon’s signed-rank test was used to compare the groups.

#### Identification of antigen-driven clonal expansions

The ALICE algorithm was used to detect clonotypes with a significantly higher number of similar TCRs than expected in the absence of clonal expansion (as predicted by the model of V(D)J-recombination) (13,14). For estimation of generative probability we run 25 iterations with 5*10^6 recombinations per iteration. Thus, the total number of simulated TCRbeta sequences (both in-frame and out-of-frame) was 1.25*10^8. Clonotypes represented by 1 UMI were excluded from the repertoires prior to analysis. A significance threshold of p<0.001 was used.

#### Association of TCRbeta motifs with HLA alleles

Incidence of a cluster was calculated as a cumulative incidence of clonotypes comprising the cluster. To identify clusters associated with HLA-B*27 or HLA-B*38 we compared occurrence of the 29 shared clusters between CD8+ SF samples of HLA-B*27+ and HLA-B*38+ patients (excluding 1 HLA-B*27+/B*38+ patient) by Fisher’s exact test with Benjamini-Hochberg adjustment.

#### Annotation with VDJdb

To annotate clonotypes with known antigen-specificity we used the VDJdb database (build 2021-02-02) (15). To exclude ambiguous sequences from the VDJdb it was filtered as follows: among clonotypes with identical CDR3 amino acid sequence specific to epitopes of several species we retained those with maximal VDJdb score, maximal number of publications and specificity to a single epitope within a species. All clonotypes with the score = 0 were excluded.

The clonotype was annotated if it had identical CDR3 amino acid sequence and V-segment with a published clonotype and the HLA allele carried by the individual matched HLA restriction of the published clonotype at 2-digit resolution.

The proportion of the T cells occupied by clones specific to EBV epitopes in paired PB and SF samples was compared by two-sided Wilcoxon signed-rank test.

## Supporting information

STROBE_checklist_v4_combined

## Data Availability

Raw sequencing data obtained for the present study are deposited in the ArrayExpress database (E-MTAB-11498)

## FUNDING

This work was supported by the Russian Science Foundation grant No 20-75-00041.

## ACKNOWLEDGEMENTS

We are most grateful to all patients and medical staff for participation in this work.

## COMPETING INTERESTS

The authors declare no conflict of interest affecting the presented results. EL is a member of the Speakers bureau of Janssen. TK is a member of Speakers bureau of: Pfizer, MSD, Novartis, AbbVie, Janssen, Lilly, Celgene, JSC BIOCAD, and Novartis-Sandoz. SE is a member of the Speakers bureau of KRKKA, MCB and JSC BIOCAD. SL and IZ provide scientific advisory service for JSC BIOCAD.

## SUPLLEMENTARY FIGURES

**Supplementary Figure 1.**
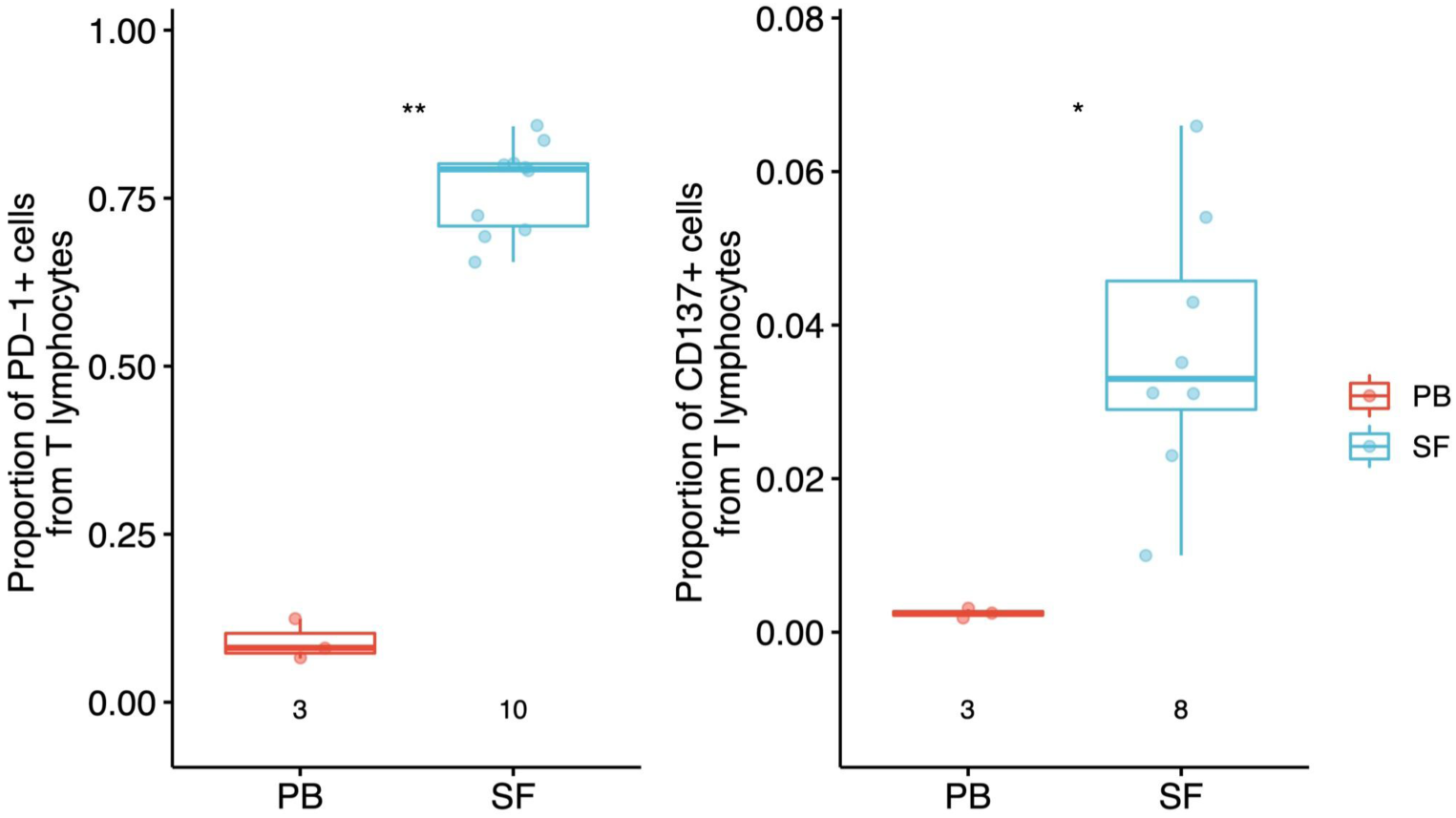
Expression of PD-1 and CD137 on synovial fluid T cells of SpA patients. Proportion of PD-1+ (left) and CD137+ (right) cells from CD3+ cells of peripheral blood (n=3) and synovial fluid (n=10 and n=8 for PD-1 and CD137, respectively) samples. Wilcoxon rank sum test: *p<0.05, **p<0.01.

**Supplementary Figure 2.**
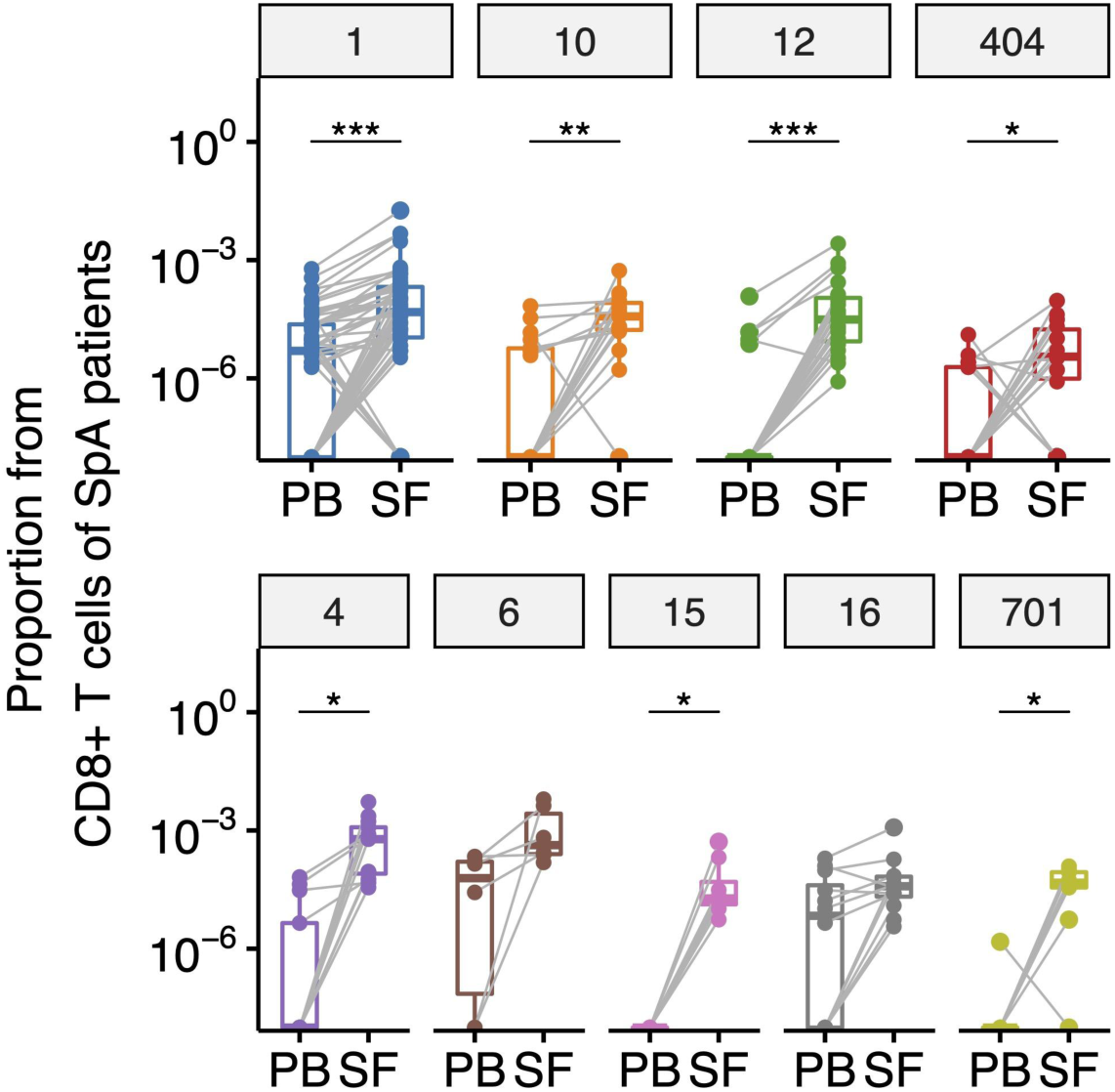
Proportion of clonotypes of identified clusters in paired PB and SF of SpA patients. T cell frequency of clonotypes comprising identified clusters in CD8+ PB and SF repertoires of HLA-B*27+ (upper, n=8) or HLA-B*38+ (bottom, n=5) SpA patients. Points represent amino acid clonotypes, and lines connect the same clonotype within a donor. Wilcoxon signed-rank test with Benjamini-Hochberg adjustment: *p<0.05, **p<0.01, ***p<0.001.

**Supplementary Figure 3.**
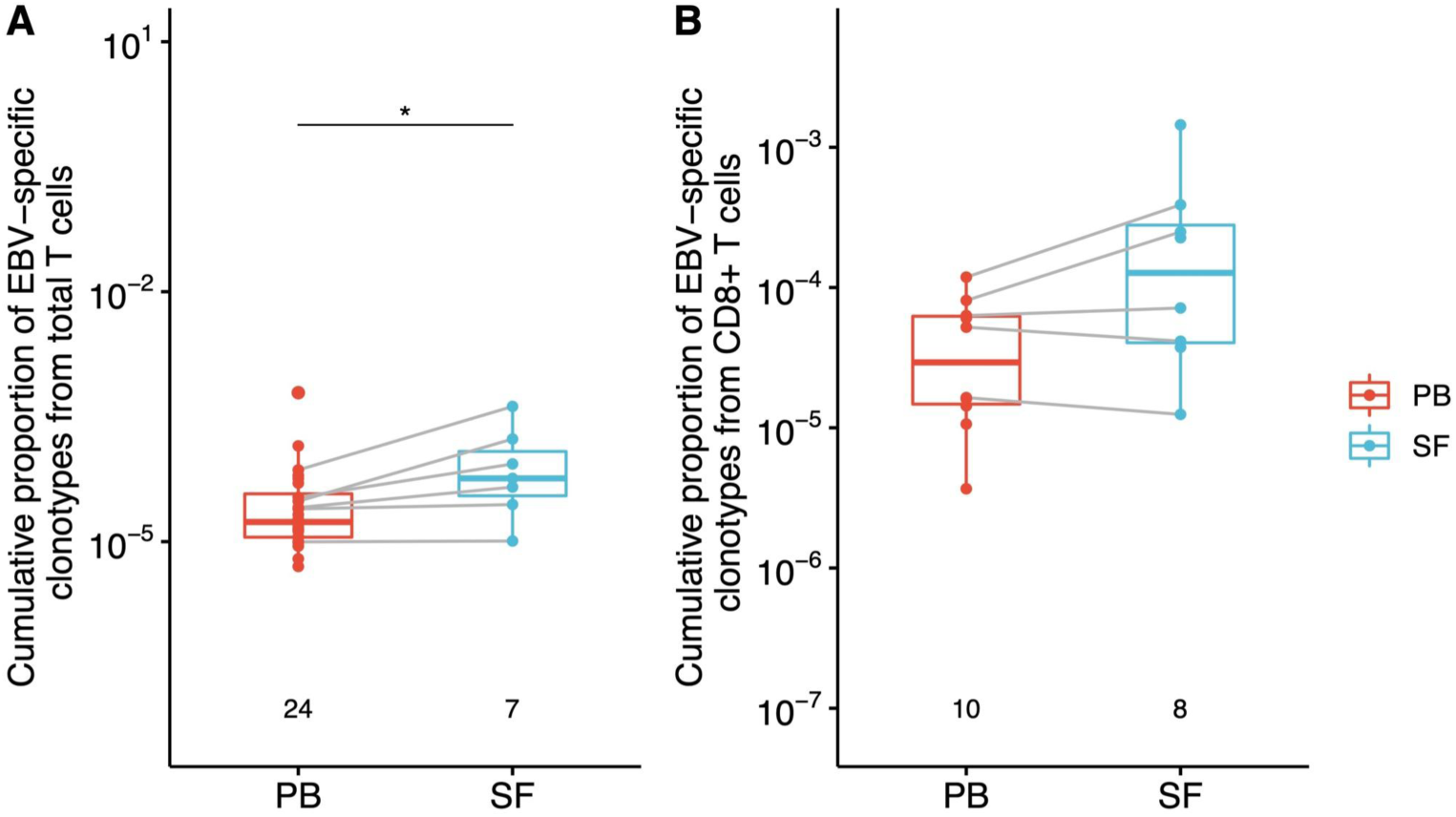
Proportion of EBV-associated T cell clonotypes in SF of SpA patients. Cumulative T cell frequency of clonotypes assigned as specific to EBV epitopes by VDJdb in total (**A**) and CD8+ (**B**) T cell repertoires of SpA patients. Lines connect paired PB and SF samples of the same donor. * Wilcoxon signed-rank test p<0.05.

## SUPPLEMENTARY TABLES

**Supplementary Table 1.**
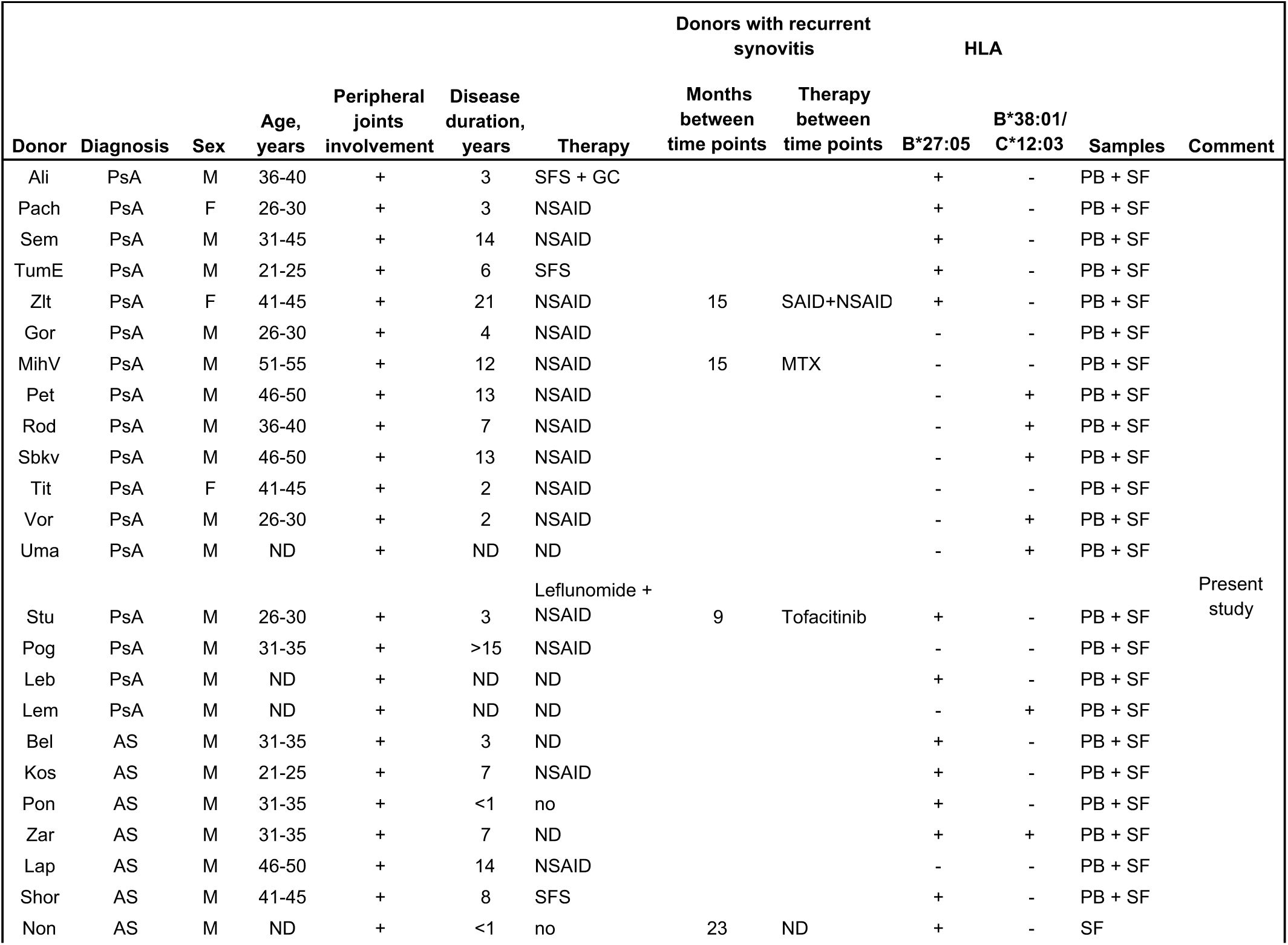
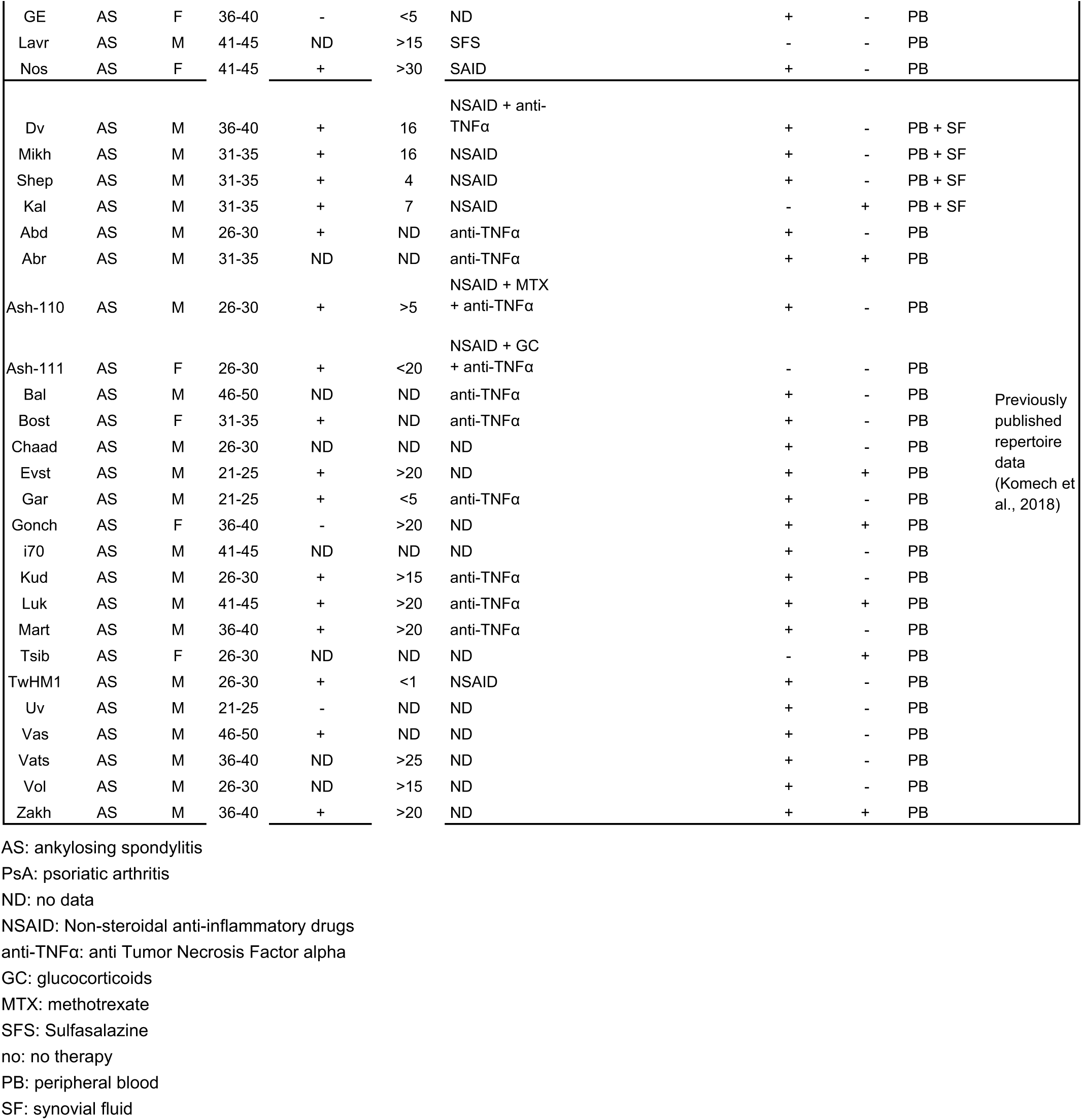
Characteristics of SpA patients.

**Supplementary Table 2.** TCRbeta repertoire profiling statistics.

| **Source** |  | **PB** | **SF** |
| --- | --- | --- | --- |
| **Number of samples\*** |  | 91 | 75 |
| **Total reads** | mean | 4325638 | 2697490 |
|  | median | 3077413 | 2799685 |
|  | q1 | 2570825 | 2048384 |
|  | q3 | 5182962 | 3293373 |
| **Total UMIs\*\*** | mean | 555404 | 440384 |
|  | median | 404166 | 284652 |
|  | q1 | 208939 | 125745 |
|  | q3 | 719048 | 556059 |
| **UMIs covered with at least 2 reads** | mean | 355661 | 256141 |
|  | median | 267258 | 207539 |
|  | q1 | 144492 | 83554 |
|  | q3 | 483323 | 358229 |
| **Total clonotypes** | mean | 141875 | 28594 |
|  | median | 97706 | 20095 |
|  | q1 | 54112 | 11372 |
|  | q3 | 201116 | 37628 |

\* Total, CD4+ and CD8+ samples cumulatively
\*\*UMI - unique molecular identifier

**Supplementary Table 3.** Occurrence of clusters shared by at least 3 donors in CD8+ SF repertoires of HLA-B*27+ and HLA-B*38+ SpA patients.

| Cluster | Total donors<br>(n=27)* | B*27+<br>donors<br>(n=15)* | B*38+<br>donors<br>(n=7)* | Fisher's<br>exact test<br>p-value | FDR-adjusted p-<br>value | Annotation | B*27+B*38+<br>donor (n=1) |
| --- | --- | --- | --- | --- | --- | --- | --- |
| 1 | 14 | 14 | 0 | **4,69E-05** | **1,36E-03** | B27-associated | 1 |
| 2 | 16 | 10 | 3 | 3,76E-01 | 7,27E-01 |  | 1 |
| 3 | 13 | 8 | 4 | 1,00E+00 | 1,00E+00 | A02-EBV | 1 |
| 4 | 7 | 0 | 6 | **9,38E-05** | **1,36E-03** | B38-associated | 0 |
| 5 | 21 | 13 | 6 | 1,00E+00 | 1,00E+00 |  | 1 |
| 6 | 6 | 1 | 5 | **4,32E-03** | **1,72E-02** | B38-associated | 0 |
| 7 | 10 | 7 | 2 | 6,48E-01 | 9,40E-01 | A02-EBV | 0 |
| 8 | 20 | 12 | 6 | 1,00E+00 | 1,00E+00 |  | 1 |
| 9 | 11 | 9 | 2 | 3,61E-01 | 7,27E-01 | A02-Influenza A | 1 |
| 10 | 10 | 10 | 0 | **5,35E-03** | **1,72E-02** | B27-associated | 1 |
| 11 | 8 | 8 | 0 | **2,25E-02** | 5,93E-02 |  | 0 |
| 12 | 11 | 11 | 0 | **3,87E-03** | **1,72E-02** | B27-associated | 0 |
| 13 | 17 | 9 | 7 | 1,21E-01 | 2,70E-01 |  | 1 |
| 14 | 5 | 3 | 2 | 1,00E+00 | 1,00E+00 | A02-EBV | 0 |
| 15 | 4 | 0 | 4 | **4,78E-03** | **1,72E-02** | B38-associated | 1 |
| 16 | 5 | 0 | 5 | **7,97E-04** | **7,70E-03** | B38-associated | 1 |
| 21 | 21 | 12 | 7 | 5,23E-01 | 9,40E-01 |  | 1 |
| 24 | 8 | 4 | 3 | 6,30E-01 | 9,40E-01 |  | 0 |
| 25 | 5 | 2 | 1 | 1,00E+00 | 1,00E+00 |  | 0 |
| 29 | 12 | 7 | 4 | 1,00E+00 | 1,00E+00 |  | 0 |
| 31 | 5 | 2 | 2 | 5,65E-01 | 9,40E-01 |  | 0 |
| 43 | 8 | 8 | 0 | **2,25E-02** | 5,93E-02 |  | 0 |
| 50 | 17 | 10 | 5 | 1,00E+00 | 1,00E+00 |  | 0 |
| 51 | 13 | 9 | 4 | 1,00E+00 | 1,00E+00 |  | 0 |
| 52 | 9 | 6 | 2 | 1,00E+00 | 1,00E+00 | A02-EBV | 0 |
| 309 | 8 | 3 | 5 | 5,24E-02 | 1,27E-01 |  | 0 |
| 404 | 11 | 10 | 0 | **5,35E-03** | **1,72E-02** | B27-associated | 0 |
| 701 | 6 | 1 | 5 | **4,32E-03** | **1,72E-02** | B38-associated | 1 |
| 1346 | 11 | 7 | 2 | 6,48E-01 | 9,40E-01 |  | 0 |

\* one B\*27+B\*38+ donor is excluded

**Supplementary Table 4.**
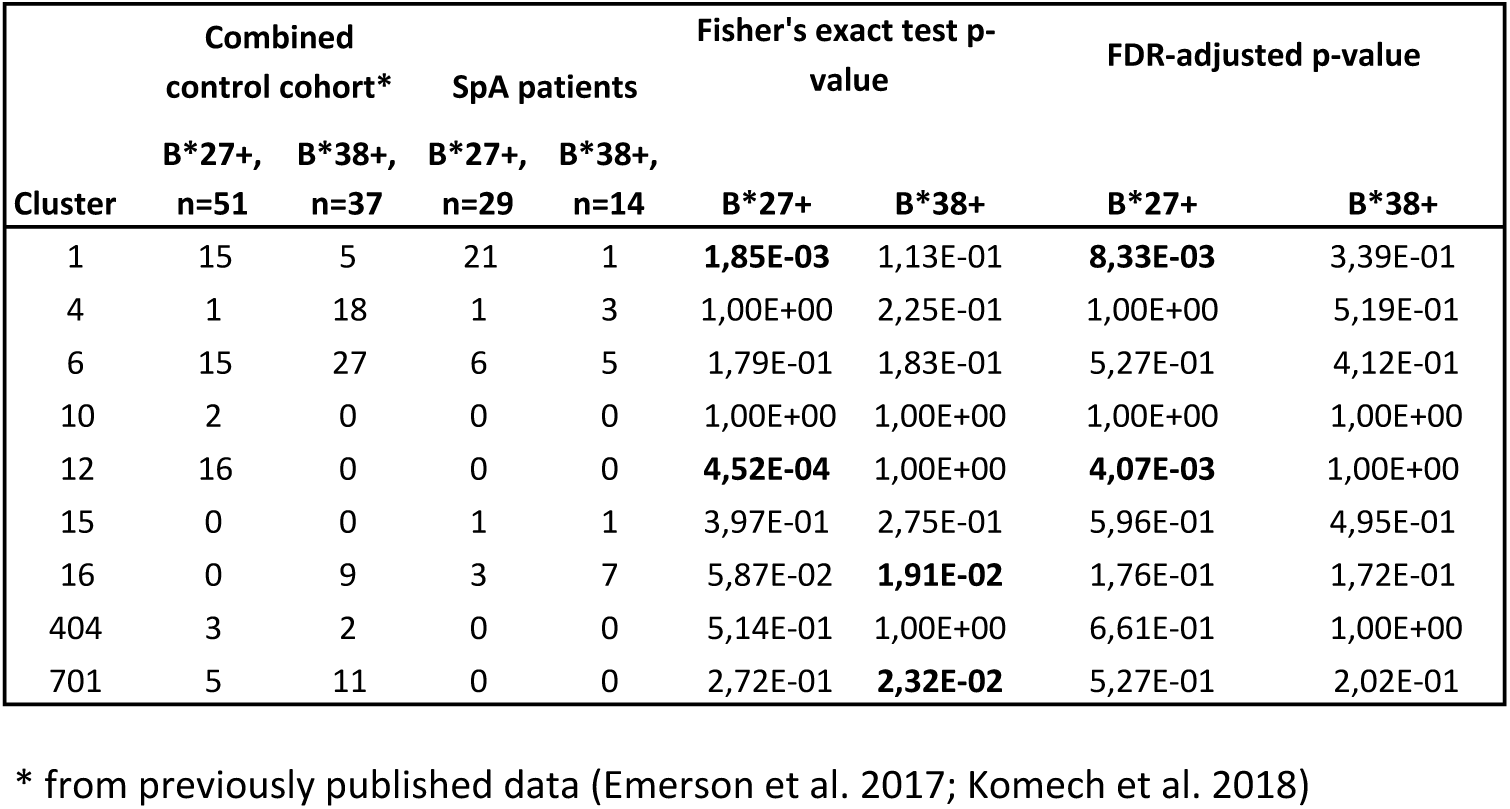
Occurrence of selected clusters in total PB repertoires of SpA patients and healthy donors.

| Cluster | Combined control cohort* |  | SpA patients |  | Fisher's exact test p-value |  | FDR-adjusted p-value |  |
| --- | --- | --- | --- | --- | --- | --- | --- | --- |
|  | B*27+, | B*38+, | B*27+, | B*38+, | B*27+ | B*38+ | B*27+ | B*38+ |
|  | n=51 | n=37 | n=29 | n=14 |  |  |  |  |
| 1 | 15 | 5 | 21 | 1 | **1,85E-03** | 1,13E-01 | **8,33E-03** | 3,39E-01 |
| 4 | 1 | 18 | 1 | 3 | 1,00E+00 | 2,25E-01 | 1,00E+00 | 5,19E-01 |
| 6 | 15 | 27 | 6 | 5 | 1,79E-01 | 1,83E-01 | 5,27E-01 | 4,12E-01 |
| 10 | 2 | 0 | 0 | 0 | 1,00E+00 | 1,00E+00 | 1,00E+00 | 1,00E+00 |
| 12 | 16 | 0 | 0 | 0 | **4,52E-04** | 1,00E+00 | **4,07E-03** | 1,00E+00 |
| 15 | 0 | 0 | 1 | 1 | 3,97E-01 | 2,75E-01 | 5,96E-01 | 4,95E-01 |
| 16 | 0 | 9 | 3 | 7 | 5,87E-02 | **1,91E-02** | 1,76E-01 | 1,72E-01 |
| 404 | 3 | 2 | 0 | 0 | 5,14E-01 | 1,00E+00 | 6,61E-01 | 1,00E+00 |
| 701 | 5 | 11 | 0 | 0 | 2,72E-01 | **2,32E-02** | 5,27E-01 | 2,02E-01 |

\* from previously published data (Emerson et al. 2017; Komech et al. 2018)

**Supplementary Table 5.**
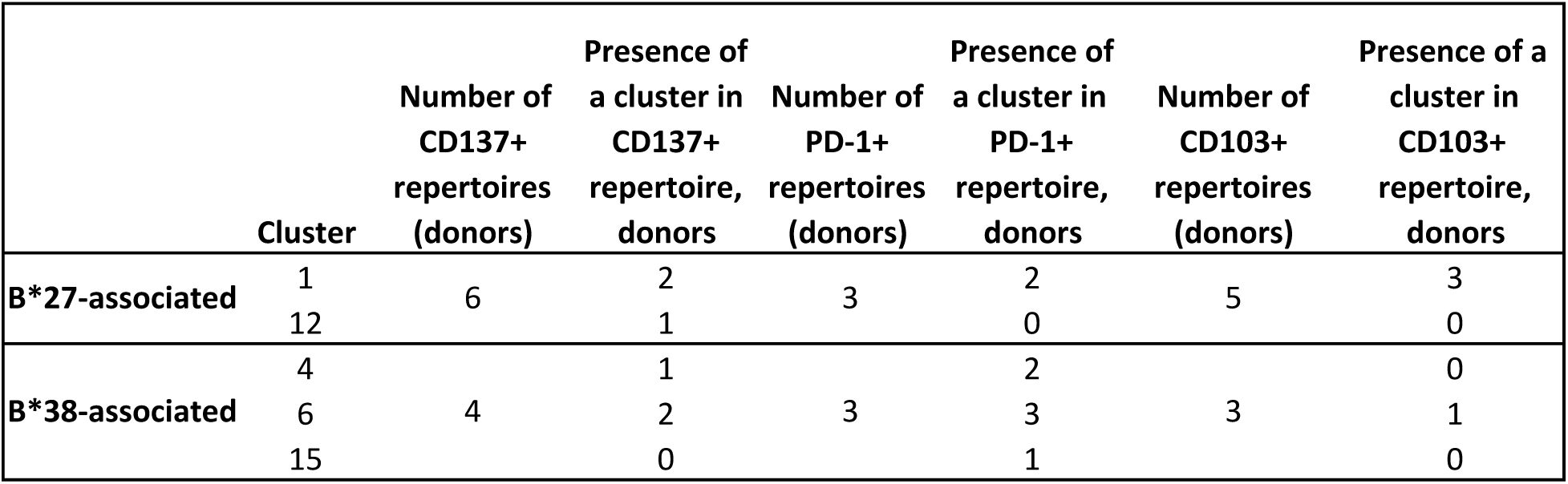
Occurrence of clonotypes of the identified clusters in PD-1+ and CD137+ repertoires of SF T cells of SpA patients.

## Notes

### Author Declarations

The study was approved by the Ethics Committee of Pirogov Russian National Research Medical University (abstract #170 18 Dec 2017)

